# “…but I know something’s not right here”: Exploring the diagnosis and disclosure experiences of persons living with ALS

**DOI:** 10.1101/2024.03.14.24304312

**Authors:** Kathleen M. Foldvari, Paul Stolee, Elena Neiterman, Veronique Boscart, Catherine Tong

## Abstract

**Background:** Amyotrophic Lateral Sclerosis (ALS), an incurable motor neuron disease, primarily affects those between the ages of 60-79, and has an approximate post-diagnosis life--expectancy of only two to five years. The condition has an unpredictable but ultimately terminal trajectory that poses a number of challenges for patients, caregivers and healthcare providers. While the diagnosis and disclosure are critical periods for intervention and support, knowledge regarding the relational, communicational and psychodynamic forces that occur within the process of diagnostic disclosure is relatively limited.

**Objectives:** The purpose of this study was to explore the experiences of persons living with ALS in the diagnosis and disclosure of the condition, and the experiences of their caregivers.

**Methods:** We conducted a focus group and in-depth individual interviews with Canadians living with ALS (n = 9), family caregivers (n = 7), a professional caregiver (n = 1), and one past caregiver (1). The interviews were transcribed, cleaned, and anonymized, and then entered into NVivo 11 for thematic analysis.

**Results:** Participants discussed the diagnosis process, including the inklings and subtle changes prior to diagnosis, attempts at self-diagnosis, and the lengthy assessment process. Time was also a consideration in the disclosure process, in which participants shared how the diagnosis disclosure was the product of longstanding conversations with their care providers. It was described as rarely a shock to finally have confirmation. Additionally, participants shared their information seeking strategies and needs for a diagnosis that, for them, typically came with insufficient information on the disease, prognosis, and next steps.

**Significance:** This project serves as an initial step in bridging the relevant gaps in our knowledge and understanding towards improved patient-centered care practices in the diagnosis and disclosure of ALS.

## Introduction

ALS is a progressive and incurable multi-system disorder characterized by muscle wasting, severe disability, and unpredictable illness trajectories. Its symptoms are caused by rapid brain and spinal cord nerve cell degeneration, and typically lead to death within three years of initial symptom onset [1-3]. ALS is a burdensome and typically terminal disease that causes great stress and uncertainty for the individual with the disease, and all those in their circle of care [4-6].

At present, there are approximately 200,000 people globally who are living with ALS [7], and it is predicted that the global incidence of ALS will increase nearly 70% by the year 2040 [8]. This anticipated increase in diagnosed cases of ALS has been attributed, largely, to the aging of the population. The number of older adults, worldwide, is expected to reach two billion by 2050 [9]; ALS is classified as an “age-dependent neurodegenerative disease” [10]. The ALS rate of incidence peaks around the eighth decade of life, while the majority of those living with the condition are between the ages of 60-79 years [8,10-12].

A lack of familiarity with the unique nature of ALS limits healthcare providers in being able to answer patients’ questions and direct their care appropriately. This lack of knowledge about the condition has the potential to create gaps in the continuity of care starting with diagnosis [2,13-15]. ALS’ relentless attack on the body’s physical functioning, as well as its incurable nature, make the disease an unfavorable diagnosis. People are reported to respond to hearing a diagnosis of ALS with a range of emotions. It is not unusual for patients to experience initial shock and distress, anger or worry [16-19]. As is true for many people faced with bad news about their health, it is common for people to struggle emotionally. Experiences with denial or wishful thinking throughout the disease course are also common reactions that are recognized in the literature [20-23]. Some studies suggest that more attention ought to be paid to patients’ emotional coping needs, as their emotional coping can have a significant impact on how they are able to engage in health planning related to their overall health outcomes [16,18,21,24,25]. In response to such evidence, general guidelines for diagnostic disclosure have been developed and implemented to help healthcare providers effectively deliver bad news to their patients, while identifying and dealing with unfavoured coping mechanisms such as denial, e.g., [14,17,18]. Though well received, many of the current guidelines were not developed or tested specifically for use in ALS care. When it comes to communication and relational care, current practices in ALS care are, and have typically been, guided by parameters outlined for cancer care focused initiatives [20,26-28].

### The diagnosis of ALS

Diagnosing ALS is typically a very involved process, as there is no one test that can determine, with certainty, whether or not the presented symptoms are related to the disease [2,13,19]. The *El Escorial* criteria for the diagnosis of ALS, originally developed in 1990 by the World Federation of Neurology and updated in 2000, are applied worldwide by clinicians and scientists in efforts to identify and classify symptoms of the disease as they are related to either upper or lower motor neuron degeneration [29].

In Canada, primary care physicians are usually a patient’s first point of contact with the healthcare system [30]. For ALS patients, and those presenting with possible symptoms of the condition, primary care services offer referral and access to multidisciplinary care clinic teams that ideally work in a coordinated way to facilitate the diagnostic process, help guide efforts in symptom management, and support the prevention of medical complications and premature death [24,30-32].

A patient’s journey often begins with complaints of preliminary symptoms such as: muscle cramps, weakness and fatigue. Shortness of breath, numbness, and/or tingling in the extremities are also common first symptoms. After noticing these first symptoms, patients will seek out a primary care physician to help make sense of their symptoms and consider their case.. The primary care physician will then make a referral to a specialized neurologist for testing and disease confirmation, if appropriate. Once ALS has been confirmed through the reasonable exclusion of other conditions, the patient’s specific needs are assessed and a care team is assembled [1,24,31,32].

A confirmed diagnosis is often a prerequisite to gaining access to specialized and funded services, such as specialized ALS clinics. A confirmed diagnosis can take upwards of a year, and misdiagnosis is relatively common in the initial stages of the process [1,2,13]. The often-convoluted process of arriving at a confirmed diagnosis of ALS can often affect patients’ relationships with their care providers, and thus their overall satisfaction with their care, making good communication an essential part of the process [1,33,34].

### The diagnosis disclosure of ALS, a critical period for intervention

While a diagnosis is defined as the “the art or act of identifying a disease from its signs and symptoms” [35], diagnostic disclosure specifically refers to the act of sharing the diagnosis with the patient and/or caregivers. Diagnostic disclosure is a term typically used when a clinician is disclosing diagnoses with serious and long-term implications, such as dementia or autism [36]. Most tools and guidelines currently available for diagnostic disclosure do not offer specific instructions for how to go about disclosing the diagnosis of ALS towards successful coping [24,26,37], although Pizzimenti and colleagues [18] do offer some guidance on the timing, location and communication of the diagnosis. Despite some existing guidance, a large number of physicians report experiencing a great deal of stress and uneasiness when breaking the news of terminal illness to patients. Meanwhile, many patients say that they are left feeling under-informed and dissatisfied after hearing the news [38-40]. There is still a need for practice-level improvements to be made to the disclosure process for ALS [38,41].

As a sensitive and stressful step in the care process, diagnostic disclosure is a critical period for intervention which is potentially predictive of future coping-related outcomes [40,42,43]. There is likely to be great benefit in working towards improving current guidelines and recommendations by expanding our knowledge base about peoples’ experiences in the process.

### Past & present approaches to diagnostic disclosure

Two commonly used tools for the diagnostic disclosure of serious conditions include the six-step SPIKES protocol, described by Baile et al. [20], and the parameters outlined by Andersen et al. [24] of the European Federation of the Neurological Societies (EFNS) task force. The SPIKES protocol lays out a series of steps to be followed during conversations in which bad news is delivered. These steps are aimed at guiding healthcare providers in gathering information, transmitting relevant information, providing support, and encouraging patients’ engagement in care planning [20]. The EFNS task force lists a number of recommendations to be considered prior to and during conversations in which a diagnosis of ALS is to be relayed. Among the recommendations listed are: a comfortable quiet setting in which to communicate the bad news; an understanding of who the patient is, and what their communicational preferences are, prior to disclosure; and the use of simple and carefully worded statements [24]. This is not dissimilar to the recommendations made by Pizzimenti et al. [18], specific to the disclosure of ALS.

Both tools (SPIKES and EFNS) have been correlated with improvements in patients’ satisfaction surrounding the manner in which the news of their illness was broken to them [38-40]. However, many physicians still report experiencing a great deal of stress and uneasiness when breaking the news of terminal illness to patients, while patients are left with many questions about their condition [38,41,44,45]. This literature suggests that parameters for diagnostic disclosure tend to be vague and do not meet expectations for supporting healthcare providers in facilitating conversations that focus on outcomes, rather than just treatment options, with their patients. This creates a risk for negative experiences in both care provision and on the receiving end [24,38,44,46,47].

A deeper understanding of varying ALS care experiences could lay the foundation for improving the diagnostic and disclosure process, potentially leading to improved coping and quality-of-life [18,38,40,41,43]. The current study seeks to understand the experiences of diagnosis and disclosure of persons living with ALS, with the support from their caregivers.

## Materials and Methods

We utilized a qualitative interview approach to capture participants’ experiences in diagnosis and disclosure of ALS [48-50]. Data collection took place in Southern Ontario, Canada, from October 2017 to June 2018.

### Eligibility & recruitment

Patients diagnosed with the various ALS types, as well as caregivers, were eligible. No participants were excluded based on their gender, socio-economic status, race, ethnicity or religious affiliation. Though the median age for ALS symptom onset is about 62 years of age, most people who develop ALS are between 40 and 70 years [8,12,51]. This study thus sought to collect data from and about adults (both male and female) roughly within the indicated age range and beyond; this study was not restricted to patients living with ALS at ages conventionally defined as “older” (i.e., 65+). The eligibility of caregivers to participate in the study was based on self-report of their involvement in the care of someone diagnosed with ALS, whether they were a partner, family member, or friend.

This study received ethics clearance from the University of Waterloo Office of Research Ethics [ORE # 22512]. In the role of a gatekeeper [52], the ALS Society of Canada shared our recruitment materials (emails and recruitment posters) with potentially eligible individuals.

### Data collection

In October 2017, a group of three patients living with ALS and six caregivers participated in an audio-recorded focus group interview in which participants discussed their experiences with diagnosis and disclosure. The focus group participants also helped develop the interview guides for subsequent individual interviews; these included one interview guide for use with patients and a second for use with caregivers.

In the Spring of 2018, thirteen participants were recruited to participate in semi-structured interviews [48]; this included four individuals who had also participated in the initial focus group. Between the focus group and individual interviews, a total of 18 unique individuals participated in this study. Interviews took place at the time and location of the participants’ choosing and were conducted over the telephone or in-person. Individual and dyad interviews were conducted based on the indicated preferences of the participants. Upon receipt of written informed consent, interviews were audio-recorded, and later were manually transcribed verbatim by the lead author. In the interviews, participants discussed their diagnosis and diagnostic disclosure experiences and provided demographic information. The lead author also composed field notes throughout the data collection period (October 16, 2017 to June 20, 2018). This study was limited in time and scope, and saturation or “data adequacy” [53] may not have been achieved.

### Description of sample

The average age of participants with ALS was 55 years, ranging between approximately 30 to 70 years (rounded for anonymity). The average age of all caregiver participants was 44 years, ranging between approximately 25 and 60 years (rounded for anonymity). The average age of caregivers, as stated, represents all types of caregiver participants in the sample excepting one family caregiver who declined to complete the demographics form. All participants lived in southern Ontario, except one who lived in British Columbia and completed their interview remotely.

The majority of the participants with ALS or similar diagnosis were male, while the majority of the caregiver participants (including past and professional) identified as female. Participants reported diagnoses of various types and motor neuron disease (MND) sub-types including probable/ atypical ALS, confirmed ALS, confirmed PLS (one person with Partial Lateral Sclerosis (PLS), a motor neuron disease similar to ALS, participated in the focus group interview), confirmed ALS – slow progressing, with some overlap between categories, where participants were given more than one related diagnosis. The overlap is attributed to initial misdiagnoses and/or multiple opinions from different healthcare providers. Participants spoke to their unique experiences identifying with the ALS diagnosis and its sub-types as they received opinions from various healthcare providers. These diagnoses were reported to have occurred between the years of 2010 to 2017. All participants described a path to diagnosis that began with either a visit to a family doctor or walk-in clinic physician upon symptom onset, followed by a referral to an ALS clinic for specialized diagnostics and care Final ALS diagnoses were made by specialized neurologists. See Tables 1 and 2 for participants’ summaries; summaries edited for confidentiality.

**Table 1.**
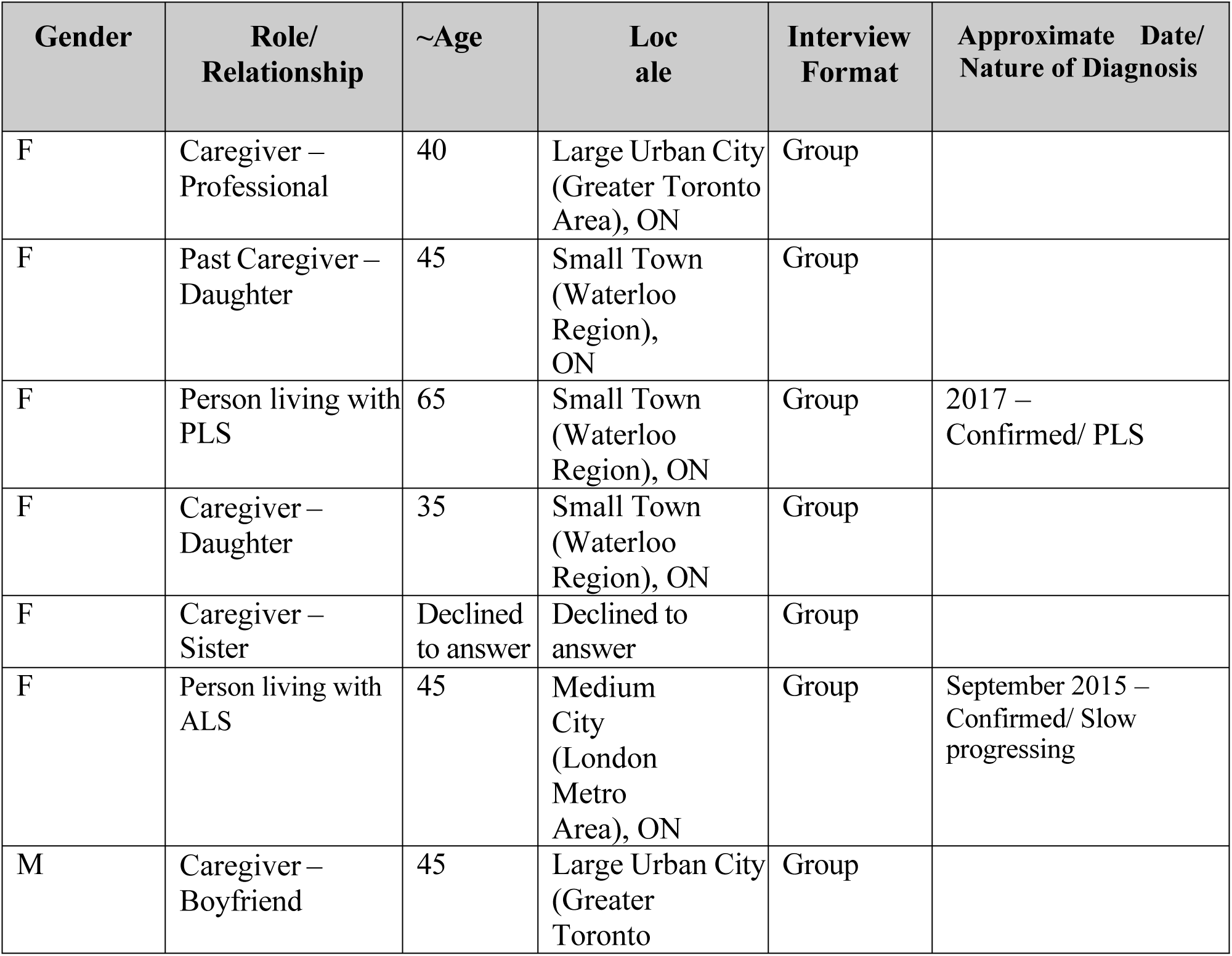
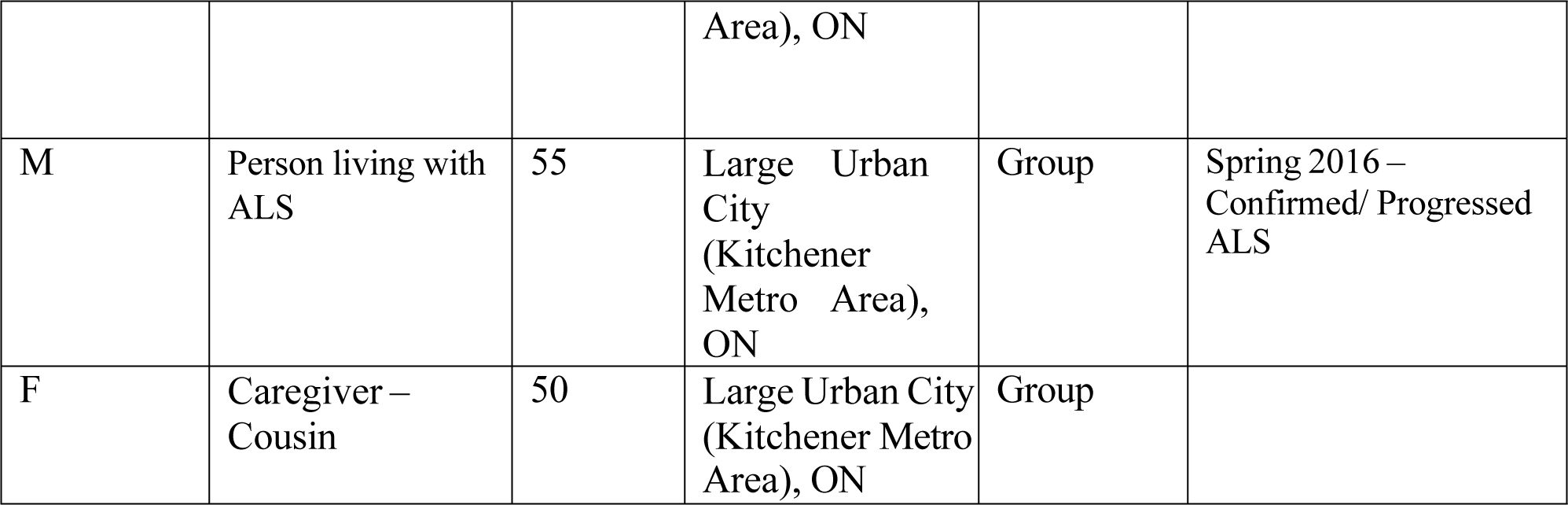
Participant Sample Summary (Focus Group Interview), n=9.

**Table 2.** Participant Sample Summary (Individual and Dyad Interviews), n=13.

| Gender | Role/ Relationship | ~Age | Locale | Interview Format | Approximate Date/ Nature of Diagnosis |
| --- | --- | --- | --- | --- | --- |
| M | Person living with ALS | 65 | Large Urban City (Kitchener Metro Area), ON | Individual | October 2016 – Probable/ Atypical |
| M | Person living with ALS | 30 | Large Urban City (Greater Vancouver Area), BC | Individual | May 2016 – Confirmed/ Limb onset |
| F | Person living with ALS | 55 | Large Urban City (Kitchener Metro Area), ON | Dyad | 2015/2016 – Confirmed |
| M | Caregiver – Son | 25 | Large Urban City (Kitchener Metro Area), ON | Dyad |  |
| M | Person living with ALS | 60 | Large Urban City (Greater Toronto Area), ON | Individual | 2015 – Confirmed/ Atypical; Lower MND Variant |
| M | Person living with ALS | 50 | Large Urban City (Kitchener Metro Area), ON | Dyad | Spring 2010 – Confirmed/ Slow progressing |
| F | Caregiver – Wife | 50 | Large Urban City (Kitchener Metro Area), ON | Dyad |  |
| F | Caregiver – Wife | 60 | Large Urban City (Kitchener Metro Area), ON | Individual |  |
| F | Person living with ALS | 70 | Large Urban City (Kitchener Metro Area), ON | Individual | 2010 – Confirmed/ Slow progressing |
| F | Person living with ALS | 45 | Medium City (London Metro Area), ON | Dyad | September 2015 – Confirmed/ Slow progressing |
| M | Caregiver – Boyfriend | 45 | Large Urban City (Greater Toronto Area), ON | Dyad |  |
| M | Person living with ALS | 55 | Large Urban City (Kitchener Metro Area), ON | Dyad | Spring 2016 – Confirmed/ Progressed ALS |
| F | Caregiver – Cousin | 55 | Large Urban City (Kitchener Metro Area), ON | Dyad |  |

### Analysis

Data were cleaned, anonymized, and then uploaded to NVivo 11 for analysis. A thematic analysis, as outlined by Braun & Clarke [54] was used. The thematic analysis conducted for the purposes of this project aimed to understand the experiences of persons with ALS through the eyes of patients and their caregivers and took place under epistemological assumptions aligned with the constructionist paradigm. Under this system of belief, themes were generated through an analysis of participant-described events and incidents, with the understanding that people build their own realities as they navigate through, and create meaning from, the world around them. The constructionist paradigm recognizes and embraces the complexity of human life, which provided a suitable basis for understanding people in their attempt to cope with a terminal illness: ‘what they do’ and ‘how they do it’ [54,55]. The data analysis process occurred inductively, meaning that the researcher did not attempt to fit the data into an existing framework or hypothesis, but rather let the data ‘tell the story’ [54].

Our analysis process included the lead author reading the transcripts twice, specifically looking for patterns in the second reading; identifying codes that represented the data, and then using printed copies of the codes to visually organize the codes thematically; defining each code, and sharing these code definitions with co-authors for review; additional readings of the transcripts and line-by-line coding in NVivo 11, and then isolating the most significant themes. These themes were then shared with the study participants. All participants were presented with summaries of the themes and asked to confirm or refute the findings.

### Strategies for rigor

Strategies for rigor [56] included member checking to verify research findings, research team debriefing, triangulating data and themes from different participant groups (e.g., caregivers and persons living with ALS), and keeping detailed field notes during data collection. Regular notetaking was used by the lead author to reflect on her perceptions, preconceptions, beliefs and values, and to attempt to separate her personal experiences with ALS from those of the participants.

## Results

Findings are clustered into two overarching themes, the first on participants’ path to diagnosis, and the second on participants’ diagnosis disclosure experience, and accompanying needs. In both themes, time and lengthy intervals were noted.

### The path to diagnosis: inklings and assessments

Participants disclosed that their path through diagnosis was initiated by changes in functional capacity or unusual sensations. For example, one participant with ALS said:

> “Before [I was diagnosed] it was about my blood sugar…The family doctor…was sending me for measurements of my blood sugar…one day…I was trying to do push-ups…and I couldn’t do it. I couldn’t do it anymore…so, that was when I went to [my family doctor]…” — [Person living with ALSI-1]

Participants described a series of events triggered by their initial symptoms that involved a visit to either their family doctor or a doctor at a walk-in clinic. It was indicated that this visit instigated initial tests ordered by the doctor, and processes of theory generating and self-diagnosis. These processes are reflected in the following statement made by a participant with ALS:

> “…I started to feel like I was being electrocuted in my feet and legs… it just went from my toes to my knees. And then as time continued to pass…[it was] moving up my body… I pushed the doctor… to give me a possibility of what it could be because I was getting a little bit frustrated. And…my wife and I had been reading tens of dozens of articles on the Internet; and gone back and forth to each other trying to cross-reference my symptoms…we came up with all sorts of things…”

> — [Person living with ALSI-5]

Both patients and doctors seemed to have been speculating as to what might be causing the symptoms, as illustrated by the statement above, as well as the following remarks made by a person living with ALS and a caregiver, respectively:

> “I was a…typical, healthy guy and noticed some atrophy in my left hand. Specifically in the muscles that control the thumb…I was a cliché quote, unquote, muscly kind of dude…I assumed that I pinched a nerve in my arm while exercising or something like that…” — [Person living with ALSI-2]

> “.. initially, the degree of weakness was so mild…that’s why I thought that it would be something else…in terms of…weight loss…they were just thinking his nutritional status was not very good…[that] he wasn’t getting enough protein…Then they [thought]…maybe other conditions also cause weight loss symptoms…”— [CAR1-8]

The conversations surrounding the first symptoms presented to participants’ primary care physicians (or first points of contact) involved a lot of back and forth in testing, and referral to various specialists, otherwise characterized as a lengthy process of trial and error. As one participant with ALS indicated: “…it was a bit of a revolving door…of characters who sort of confirmed my diagnosis…people in and out…” [Person living with ALSI-2]. In some cases, misdiagnosis and ineffective surgery or treatment aimed at symptom management were also experienced. One participant with ALS said: “[It was] all just sort of symptom management while we were trying to figure out what the problem was and once targeted we could treat it…hopefully” [Person living with ALSI-5].

Others stated:

> “…my hands started getting weak. I could barely turn the keys to unlock my car. So, I went to the doctor and they said, something’s not right… And then…[the doctor] said: “I think you have carpal tunnel”. So they did the surgery on both hands and that didn’t really help…Then that doctor said it must be something different…” — [Person living with ALSI-9]

> “I was a PSW [personal support worker] and I fell in the bathtub…it was drop foot…and I knew something wasn’t right…When I fell in that bathtub, I was fine. I could run and jump until I fell in that bathtub…I kept working…and then I went to [the doctor]…and I was getting acupuncture. For many years I kept going and trying different things all the time…” — [Person living with ALSD-3]

The initial back and forth was ultimately followed by a determination, through a process of elimination, that the presented symptoms were most likely neurological in nature. This is said to have prompted final referrals made either to first, a local neurology specialist, or directly to a major city clinic neurologist for similar assessments and observations of symptom progression. One participant with ALS asserted*: “…basically they rule everything out, and then as you continue to get worse, they say: “Hey, you got ALS””* [Person living with ALSI-2]. Two other participants explain similar experiences:

> “…it’s not an obvious thing, like you’re going to break a leg, where you go into any ER and they put a cast on you, and you’re out the door type of thing. When it is a diagnosis of exclusion and you don’t know what it is…you got to figure out what it is or what it isn’t” — [Person living with ALSI-5]

> “…they try to rule out everything else. Then they say you probably have ALS. But, [they] refer [me] to a team in [a major city]…and they basically did the same tests, and said the same thing: that I probably have ALS. Then they said about nine months later they would follow-up…and then they said: “okay, you have ALS”. — [Person living with ALSD-6]

This led some directly to acceptance, while others sought a second opinion first:

> “…I went to [the clinic]…I [told] them, I don’t know what I have. I don’t know if I have ALS or what I have. So, I asked for a second opinion and they said I had a neurological disease. So I said, I don’t know what I have, but I know something’s not right here” — [Person living with ALSD-3]

Overall, the process of diagnosis was described by participants as one characterized by a great deal of back and forth, frustration, and hopeful theorizing. Participants indicated putting a great deal of effort into ascertaining a label that best suited the symptoms that they were experiencing.

### The diagnostic disclosure process: intervals and information

For the participants in this study, all conversations surrounding diagnosis and prognosis were conducted in-person, regardless of the healthcare provider that was relaying the information. These conversations were indicated to have typically gone on for nine months to about a year. In two cases, a confirmed diagnosis was reached after two to three years:

> “…there was maybe a year…maybe nine months of activity before [I received a confirmed diagnosis]…but that’s the day essentially, that they exhausted all of treatments and were monitoring my progression. Then they essentially concluded and voiced to me that I had ALS” — [Person living with ALSI-2]

> “I think it wasn’t until [three years later] that we were really told: “Okay, at this point between three different hospitals, we have run every conceivable test…and in the absence of finding anything else that we could pin this on we have to conclude that you have lower motor neuron onset ALS”” — [Person living with ALSI-5]

On the other hand, a confirmed diagnosis was reached after a matter of weeks for another case.

The process of receiving a confirmed diagnosis was described as not coming as a surprise, but as confirming the suspected, based on the patients’ own research. This is reflected in these quotes from persons living with ALS:

> “…and between the first neurologist and the second I had done a lot of research, so I had already decided that’s what I had. So, it wasn’t a surprise” [Person living with ALSI-5].

> “I was hoping for good news, but at the same time I wasn’t completely surprised about the bad news…given the independent study I did….I’ve always had terrible luck. So right off the bat and given me reading the Wikipedia page and noticing the physical symptoms I sort of concluded independently…and with my pessimism…that I had ALS. But yeah, I was noticing continued muscle atrophy, continued weakness, gradual, but obvious, over those nine months. And when I finally got the word what it was, it was one of these things that I had been expecting to hear I think” — [Person living with ALSI-2]

After receiving a confirmed diagnosis, participants described a subsequent process of convening a multidisciplinary care team. For example, one person living with ALS says: *“…once I got a confirmed diagnosis from the specialized ALS clinic, it’s been a whole care team of a half dozen individuals that I see quite regularly”* [Person living with ALSI-2]. Participants perceived their concrete diagnosis as pivotal, such that it sparked a will to push forward and reassess personal priorities, such as for this participant who declared: *“I can’t go anywhere…I have too much to live for!”* [Person living with ALSD-3].

Similarly, this person living with ALS shared:

> “So essentially what happened…from that day forward it’s like: okay, well, you have three to five years or whatever math they give you….and pardon my language, but it’s like: fuck that! I’m not going in the office anymore. You know what I mean? Like what’s in it for me?” — [Person living with ALSI-2]

In trying to digest the diagnostic and prognostic information that was presented to them, participants said that they took it upon themselves to educate themselves about their condition:

> “So I told [my wife] that night when I got home from work. And…when we went to bed, we started looking it up on the Internet. And that’s how we realized that’s all we had for those six weeks [until our next appointment]” — [Person living with ALSD-6]

Participants criticized the healthcare system for long waits between appointment times and complained that one of their most significant struggles in the diagnostic disclosure process was the limited amount of information provided about the condition they were just diagnosed with. One person living with ALS stated: *“…I knew what I had when I went to the first neurologist…when she told me I probably have ALS. But, I didn’t even know what that was, and she wouldn’t tell me”* [Person living with ALSD-6]. In this case, the participant expected to receive information and education about their condition from their neurologist. Instead, the neurologist gave the person pamphlets and told the person to “go home and look it up on the Internet”. Their caregiver added: *“…when you first read [the pamphlets they give you]…it’s sickening. You just read what it is…you don’t know. Just to be thrown [the information], with no lead up to it is awful”* [CARD-7]. This scenario highlights the lack of familiarity and discomfort with ALS diagnosis disclosure and its direct impact on patient care.

Participants discussed that having a concrete diagnosis was helpful, in that it granted them access to support from social organizations such as local ALS Society of Canada chapters and home and community care services, and eligibility to participate in treatment trials and clinical studies:

> “I am going to be trying that new medicine…hopefully in the next few weeks…[through the ALS clinic]…So I was waiting for the phone call today…[from the ALS clinic neurologist]…” — [Person living with ALSD-3]

> “…we filled out a million forms…So I’m now on the wait list for an interview for…direct funding. And I now have…through [the local health authority], doing personal support hours one hour daily. Which basically is: come in make me a coffee, shower me…Cool. I’m hooked up with [the local hospice], but I haven’t heard back from them yet…” — [Person living with ALSD-10]

Generally, experiences of diagnostic disclosure were described as not a surprise or a shock. Participants described having been grateful to be through the diagnosis process, such that they were then able to begin to focus on establishing next steps and understanding care and treatment options with a consistent care team.

## Discussion

Current recommendations for diagnostic disclosure, as laid out by Baile et al. [20] and Andersen et al. [24], advise verbal in-person sharing of diagnosis and prognostic information. According to participants, this is indeed the way such conversations are typically carried out. Participants did not share evidence to suggest that healthcare providers were implementing recommendations such as offering an invitation to share information or exploring emotions as in the SPIKES protocol and EFNS task force report [20,24]. However, the general process of care as described in the literature was corroborated by the findings of this study. Persons with ALS and caregivers in the current study explained that their journeys through the care system began with the presentation of mild symptoms to a primary care physician, followed by a diagnostic process of elimination, and a referral to a neurologist, a neurology specialist, or an ALS clinic team in a major city. A formal diagnosis from a neurologist provided them access to specific supports. This path is consistent with the descriptions of multiple sources both in and outside of Canada [1,15,24,31,32]. Our findings are also consistent with the work of Hogden et al. [16] who determined that the nature of the terminal prognosis associated with ALS can make it difficult to manage dynamics in providing and receiving healthcare, as well as patients’ engagement in their care.

Most participants in our study were not surprised by their diagnosis at the point of formal disclosure. Participants indicated that after having gone through a lengthy process of trial and error, and having done their own theorizing and research throughout, receiving confirmation of their diagnosis did not come as a shock, in contrast to the findings of some prior studies. For example, Hogden et al. [16] reported that health professionals described patients as being shocked by the diagnosis of ALS and as having difficulty accepting it. These reactions were seen as delaying the provision of additional information and the commencement of care planning. The same research group studied the perspectives of patients’ experiences of receiving an ALS diagnosis, again finding that shock was a common reaction [57]. On the other hand, several patients in the latter study indicated that the diagnosis was not a surprise, but a confirmation of their own conclusions [57], which was consistent with experiences described by participants in our study.

Emotions of surprise and shock have been studied in the experiences of people receiving a cancer diagnosis. Plage and Olsen [58] studied the illness narratives of people living with cancer and found that surprise and shock (as an intense form of surprise) conveyed the implication that the diagnosis of cancer for those expressing surprise was unexpected and unpreventable and thus not the individuals’ responsibility. In this study, the experience of surprise, or the lack thereof, for persons living with ALS, reflected a different illness experience. Persons living with ALS were not surprised by the diagnosis because they had been led to that conclusion by a lengthy diagnostic process and their own information-seeking, and not because they felt some sense of responsibility for having the condition (as might be the case for a long-time smoker who is diagnosed with lung cancer).

Participants described the Internet as a significant tool in their quests to identify the cause of their symptoms. Patients and caregivers talked about doing their own research using the Internet as a tool. Participants explained that they brought the results of their searches and discussions to their healthcare providers for further consideration and conversation. The findings of Abdulla et al. [59] on information seeking behaviors on patients and their care were similar to the findings of this study, in that patients and their caregivers sought information from multiple sources, with many relying on Internet sources for information (as was also found by [60]), and that they wanted to share and discuss the information they found with their healthcare providers. Conducting their own independent research seemed to offer participants in the current study a sense of control and authority during the diagnosis process. This information seeking behavior may have contributed to their familiarity with their condition, enough so to avoid surprise when they received their formal diagnosis.

Once they received confirmation of the suspected diagnosis, participants were finally able to put a name to their condition and begin exploring their treatment options. In the case of this sample, it was most common for participants to experience more of an intense reaction to their initial symptoms, as they worried about their ability to continue to perform optimally in life and at work. Participants began to reason and rationalize increasingly as they grew more aware of the limited treatment options that existed for their condition; this was until they slowly began to make peace with their circumstances, potentially facilitated by extended rather than abrupt processes in diagnostic disclosure or confirmation.

Jakobsson Larsson and colleagues [14] found ALS patients to be less engaged in information seeking, while continuous information seeking was quite common among persons with ALS in this study. This may highlight a difference in coping needs for persons living in community and patients in hospital, a setting in which knowledgeable healthcare providers are typically more present and available. Participants described this as a means of informing their own understanding of their condition in order to curb uncertainty, given the limited and sometimes untimely information they received from healthcare providers. Participants described information provision as generally lacking in their healthcare experience. However, the sense of control and autonomy that they felt in doing their own research about their condition is consistent with the empowerment to improve disease management and decision making that Pizzimenti et al. [18], talk about fostering, using communication that considers the information needs and goals of patients, as well as social, culture and other personal factors. It also suggests that more formal direction on where and how to seek useful information may help patients engage with the diagnosis process in a way that can reduce prolonged processes in coming to a diagnosis by being referred to a specialist for symptoms mistaken for other conditions [15,18]. Though concerns of overwhelming patients with too much information, too soon, have been discussed by research groups such as McCluskey et al. [38] and Connolly et al. [39], this present study suggest that persons with ALS want more information about what to expect in their condition, and about available resources, than they are currently receiving [61]. It is understandable that a long period of diagnostic uncertainty along with worsening symptoms would motivate persons to seek information from the Internet. The well-known concerns about the limited reliability of online health information [59-61], which was acknowledged by some participants in this study, heighten the importance of timely information from healthcare providers.

As indicated by both patients and caregivers, such information would facilitate coping by providing people with a platform from which to create their own plans for proactive action towards better management of their current and anticipated needs.

### Strengths and limitations

This study used multiple types of data collection (a focus group interview, individual and dyad interviews). The focus group interview allowed for an initial exploration of themes, and the dyad and individual interview formats were principally driven by participant preference. Great care was taken to center and focus on the voice of the person living with ALS. Across these different types of data collection, the themes were largely consistent. Member checking with all participants was used to further ensure their voices and experiences were appropriately captured in the analysis.

Limitations of this study include the limited reflection of the voices of older persons living with ALS. Few persons with ALS over the age of 65 years participated in the study; some potential participants over the age of 65 were too far progressed in their disease to participate. This can be considered a limitation of the current study, as the experiences of persons with ALS potentially with the highest needs were not captured. Future research should give careful consideration to recruitment methods.

## Conclusion

This project serves as an initial step in bridging the relevant gaps in our knowledge and understanding toward improved patient-centered care practices in the diagnosis and disclosure of ALS. This study has highlighted the importance of continuing, meaningful communication between patients and healthcare providers to minimize uncertainty and to support care planning. Further research will be necessary to corroborate and extend the study results. It will be necessary to incorporate the voice of healthcare providers that play a role in supporting people living with ALS and other closely related conditions, so that barriers and resource limitations that they might face in trying to fulfill the needs and desires of their patients can be identified. Religious-, cultural-, age-, and gender-specific needs will be important areas of continued exploration. These current and future findings, together, could support and improve the diagnosis and diagnosis disclosure experiences of persons living with ALS, and their caregivers and providers.

## Data Availability

Data are contained within the manuscript.

## Acknowledgements

The authors gratefully acknowledge the persons with lived experience of ALS who participated in the interviews for this project, and the ALS Society of Canada for their assistance with recruitment. The authors also thank Veronica Sacco and Christiano Choo for their help in reviewing and editing earlier versions of this paper.

